## Supplementary material for "Diet-wide analyses for risk of colorectal cancer: prospective study of 12,250 incident cases among 543,000 women in the UK"

### Supplementary Methods

#### Assessment of dietary intakes using the dietary questionnaire

The three-year re-survey questionnaire (forming the baseline for this study) asked participants about their diet during a typical week, including 130 quantitative or semi-quantitative questions on frequency of intake of specific foods and food groups.

*Meat*

For the questions on meat, women were asked to select the types of meat they consumed about once a week or more from the following types: beef, bacon, chicken/poultry, lamb, ham, kidney, pork, sausages, liver/pâté, beefburger/hamburger. They were also asked if they never ate meat. A separate question asked women to report the total number of times meat was eaten per week. For these analyses, we determined the portion of different meat types consumed by combining the information on consumption frequency with the types of meat selected and multiplying these by standard portion sizes. We then summed intakes by groups as follows: chicken, red meat (beef, pork, lamb, kidney, liver, burger), and processed meat (sausage, bacon, ham). We also combined red and processed meat as a separate variable. Intake frequencies were grouped into fifths and we estimated trends in risk across these baseline categories.

*Fish and seafood*

For the questions on fish, women were asked to select the types of fish they consumed about once a week or more from the following types: tuna, sardines, trout, fish and chips, salmon, mackerel, kippers/herring, cod/haddock/other white fish, and other seafood. They were also asked if they never ate fish. A separate question asked women to report the total number of times fish was eaten per week. For these analyses, we determined the portion of different fish and seafood types consumed by combining the information on consumption frequency with the types of fish selected and multiplying these by standard portion sizes. We then summed intakes by groups as follows: oily fish (sardines, salmon, trout, kippers, and mackerel), and non-oily fish (seafood, fish and chips, tuna, and white fish). Participants were then divided into groups of intakes (<1, 1+ wk) and we estimated trends in risk across these baseline categories.

*Vegetables and legumes*

For the questions on vegetables and legumes, women were asked to select the types of vegetables and legumes they consumed about once a week or more from the following types: aubergine, carrots, courgettes, beetroot, parsnip, lettuce, tomatoes, swede, spinach, sweetcorn, avocado, celery, green beans, green/red peppers, mushrooms, cucumbers, broccoli, cabbage, cauliflower, brussel sprouts, onion, garlic, leeks, green peas, baked beans, chick peas/lentils, soya/tofu. Two separate questions asked women to report the total number of times cooked vegetables and salad/raw vegetables were eaten per week. To rank the women, we grouped the intake frequencies across these two questions and grouped them into the following categories: <10, 10-14, 15-19, 20-29, 30+ pw. We then estimated trends in risk across these baseline categories. We also included the 26 individual vegetable and legume items as they were asked (grouped as once or more per week versus less than once per week). Aubergine consumption was not asked about in the Oxford WebQ and was therefore excluded from this analysis.

*Fruit*

Women were asked to select the types of fruit they consumed about once a week or more from the following types: apples, bananas, oranges/satsumas, grapefruit, pears, stone fruit. We then categorized women into those who reported eating fruit less than once per week, and once or more per week. Three separate questions asked women to report the total number of times fresh fruit, dried fruit, and stewed or tinned fruit were eaten per week. For dried fruit, and stewed or tinned fruit we categorized women into those who reported eating fruit less than once per week, and once or more per week. The question on fresh fruit was not comparable to the Oxford WebQ (which asked about consumption of fresh, frozen, and tinned fruit combined) and therefore not used. To rank the women by total fruit intake (excluding juice) we grouped the intake frequencies across the fruit subtypes, dried fruit, and stewed or tinned fruit and grouped them into the following categories: ≤5, 6-9, 10-14, 15-19, 20+ pw. We then estimated trends in risk across these baseline categories.

*Milk and ice cream*

We estimated dairy and soya milk intake using the questions on cups of milk intake (including hot cocoa), type of milk consumed, bowls of breakfast cereal, cups of tea, and cups of coffee. Women were asked which types of milk or cream they drink once a week or more often. Those who selected ‘full cream’, ‘semi-skimmed’, or ‘skimmed’ were assigned dairy milk, whereas those who selected ‘soya’ were assigned soya milk. We then estimated their total daily dairy or soya milk consumption by summing their milk intakes assuming the following: 95 mL of milk for each cup of milky drinks, 100 mL of milk for each bowl of breakfast cereal, 35 mL of milk for each cup of tea or coffee. For dairy milk, participants were then divided into fifths and we estimated trends in risk across these baseline categories. For soya milk participants were categorized into low and high and analyses were conducted with high versus low. Ice cream intake was included in the question about the types of milk or cream consumed each week and categorized as high versus low.

*Wholegrains*

We estimated wholegrain intakes using questions on intakes of pasta, rice, brown/wholemeal bread, crackers/crispbread, sweet biscuits, cake/puddings, bran cereal, biscuit cereal, oat cereal, muesli, and other cereal types. We used the wholegrain content of foods consumed in the UK to calculate the grams of wholegrains in each item [1] and then totalled the gram intakes across these foods to get a total intake for each woman. Participants were then divided into fifths of intakes and we estimated trends in risk across these baseline categories. The mean intake of 42.8 g of wholegrains in the highest quintile is equivalent to the amount in 2.5 slices of wholemeal bread.

*Remaining foods and beverages*

Women were asked to report how often they consumed the remaining foods and beverages which we categorised as follows based on the distribution of the data: chips (0, 1, 2+ times pw), potatoes <3, 3, 4, 5, 6+ times pw), pasta/spaghetti (0, 1, 2+ times pw), rice (0, 1, 2+ times pw), cheese (≤1, 2, 3, 4+ times pw), eggs (≤1, 2, 3, 4+ whole eggs pw), slices/pieces of white bread (0, 1-9, 10-19, 20+ slices pw), slices/pieces of wholemeal bread (0, 1-9, 10-19, 20+ slices pw), crackers/crispbreads (0,1-6,7+ number pw), sweet biscuits (0, 1-9, 10+ number pw), dairy desserts (≤2, 3-4, 5-6, 7+ number pw), cakes/puddings/pies/buns (0, 1-2, 3+ number pw), chocolate (≤1, 2-3, 4-5, 6+ pieces pw), nuts (<1, 1, 2+ Tbs pw), soup (0, 1, 2, 3+ bowls pw), gravy/cream cheese/sauces (≤1, 2-3, 4-5, 6+ Tbs pw), crisps (0, 1, 2, 3+ packets pw), boiled sweets(0, 1, 2-6, 7+ number pw), jam (0, 1, 2, 3+ Tbs pw), breakfast cereal (<7, 7, 8+ bowlspw), alcohol (0, 1-5, 6-10, 11+ drinks pw), tea (0-1, 2-3, 4-5, 6+ cups pd), coffee (0-1, 2-3, 4-5, 6+ cups pd), water, fizzy/soft drinks (0, 1, 2+ glasses pd), fruit juice (<2, 2-4, 5-7, 8+ glasses pw), fruit squash (0, 1, 2+ glasses pd), and ice cream (<1, 1+ (portion assumed) pw). We then estimated trends in risk across these baseline categories.

*Nutrients*

We grouped women into fifths and then estimated trends in risk across these five baseline categories.

#### Measurement error correction

In this example for calcium, we show the mean intakes in the baseline dietary survey and in the first Oxford WebQ women completed by quintiles of calcium intake in the baseline dietary survey. We assigned the mean Oxford WebQ intakes in women who completed at least one Oxford WebQ to each baseline quintile for all women, and calculated a trend variable.

| Baseline dietary survey (N=542,778) | | Oxford WebQ (N=36,597) |
| --- | --- | --- |
| Categories of calcium intake | Mean calcium intake (mg/day) | Mean calcium intake (mg/day) |
| Q1 | 492 | 828 |
| Q2 | 688 | 912 |
| Q3 | 818 | 970 |
| Q4 | 959 | 1037 |
| Q5 | 1252 | 1126 |
| *Range (mg/d) Q1 to Q5* | *760* | *298* |
| HR^1^, 95% confidence intervals per 300 mg | 0.94, 0.92-0.97 | 0.83, 0.77-0.89 |

^1^Associations between calcium and colorectal cancer incidence separately using Cox proportional hazards regression models stratified by year of birth, date of completion of the 3-year re-survey (which is baseline for this study), and region of residence (10 geographical regions (9 in England), and Scotland), and adjusted for socioeconomic group (fifths, based on the Townsend deprivation score, unknown), highest educational qualification (none, technical, secondary, tertiary, unknown), body mass index (<20, 20-22.49, 22.5-24.9, 25.0-27.49, 27.5-29.9, 30-32.49, 32.5-34.9, 35+ kg/m2, unknown), height (<160, 160–164.9, ≥165 cm, unknown), strenuous exercise (none, ≤ once per week, > once per week, unknown), dietary energy intake (except for the analysis of energy and risk; fifths, unknown), alcohol (none, 1-5, 6-10, ≥ 11 drinks per week, unknown), smoking (never, past, current 1–4, current 5–9, current <10, current 10–14, current 15–19, current 20–24, current 25–29, current ≥30 cigarettes per day, unknown), current use of hormonal therapy for menopause (no, yes, unknown), and family history of bowel cancer (no, yes). ^1^ Trend uses the five re-measured intakes from the Oxford WebQ

### Supplementary Tables

### Supplementary Table 1. Average food and nutrient intakes in first Oxford WebQ (N=36,597)

| **Food or nutrient** | **Mean** | **SD** |
| --- | --- | --- |
| Cereal g/day | 69.7 | 81.4 |
| Brown bread g/day | 8.44 | 15.25 |
| White  bread g/day | 4.07 | 4.07 |
| Rice g/day | 14.4 | 43.4 |
| Crispbread g/day | 9.20 | 22.0 |
| Pasta g/day | 23.0 | 68.2 |
| Dairy milk g/day | 198 | 160 |
| Yogurt g/day | 66.3 | 80.1 |
| Cheese g/day | 20.7 | 27.7 |
| Red and processed meat g/day | 47.4 | 62.1 |
| Red meat g/day | 34.6 | 57.2 |
| Processed meat g/day | 12.8 | 28.6 |
| Chicken g/day | 29.2 | 56.2 |
| Oily fish g/day | 13.2 | 34.7 |
| Non-oily fish g/day | 16.5 | 44.0 |
| Eggs g/day | 19.7 | 41.8 |
| Soup g/day | 42.9 | 92.9 |
| Vegetables g/day | 260 | 207 |
| Potatoes g/day | 84.4 | 100.6 |
| Chips g/day | 28.9 | 72.7 |
| Fruit g/day | 249 | 185 |
| Dried fruit g/day | 9.80 | 25.66 |
| Crisps g/day | 5.0 | 13.2 |
| Nuts g/day | 6.63 | 16.3 |
| Cakes and pastries g/day | 8.60 | 23.0 |
| Sweet biscuits g/day | 6.96 | 14.7 |
| Chocolate g/day | 7.60 | 18.55 |
| Boiled sweets g/day | 2.20 | 12.1 |
| Jam g/day | 4.54 | 7.82 |
| Gravy/cream cheese sauce g/day | 7.50 | 18.2 |
| Fruit squash g/day | 31.6 | 110 |
| Fruit juice g/day | 87.0 | 138 |
| Fizzy drink g/day | 23.5 | 103 |
| Tea g/day | 535 | 356 |
| Coffee g/day | 338 | 282 |
| Alcohol g/day | 11.2 | 16.1 |
| Energy kj/day | 8270 | 2363 |
| Carbohydrates g/day | 234 | 77.0 |
| Wholegrains g/day | 34.5 | 24.9 |
| Englyst fibre  g/day | 17.1 | 7.17 |
| Total sugars g/day | 117 | 48.2 |
| Free sugars g/day | 49.9 | 32.0 |
| Total fat g/day | 77.2 | 31.2 |
| Monounsaturated fat g/day | 25.1 | 11.5 |
| Polyunsaturated fat g/day | 13.9 | 7.97 |
| Saturated fat g/day | 30.0 | 13.1 |
| Protein g/day | 78.5 | 24.5 |
| Carotene µg/day | 3713 | 3225 |
| Retinol µg/day | 547 | 1338 |
| Thiamine mg/day | 1.93 | 4.19 |
| Riboflavin mg/day | 2.03 | 0.72 |
| Vitamin B6 mg/day | 2.13 | 0.76 |
| Folate µg/day | 303 | 125 |
| Vitamin B12 µg/day | 6.87 | 5.39 |
| Vitamin C mg/day | 164 | 112 |
| Vitamin D µg/day | 2.97 | 3.37 |
| Vitamin E mg/day | 9.67 | 4.61 |
| Calcium mg/day | 980 | 360 |
| Magnesium mg/day | 339 | 103 |
| Phosphorus mg/day | 1451 | 417 |
| Potassium mg/day | 3784 | 1176 |
| Zinc mg/day | 9.52 | 3.38 |

### Supplementary Table 2. Relative risks and 95% confidence intervals for the associations of 97 dietary factors with risk of colorectal cancer, sorted by p values

| **Food or nutrient** | **Increment** | **RR (95% CI)**^1^ | **P value** |
| --- | --- | --- | --- |
| **FDR significant** |  |  |  |
| Alcohol | 20 g/day | 1.15 (1.09,1.20) | 0.00000001 |
| Calcium | 300 mg/day | 0.83 (0.77,0.89) | 0.00000014 |
| Dairy milk | 200 g/day | 0.86 (0.81,0.92) | 0.00000777 |
| Phosphorus | 300 mg/day | 0.84 (0.78,0.91) | 0.00001196 |
| Riboflavin | 1 mg/day | 0.83 (0.75,0.91) | 0.00016233 |
| Magnesium | 100 mg/day | 0.84 (0.77,0.92) | 0.00016893 |
| Wholegrains | 20 g/day | 0.90 (0.85,0.95) | 0.00018866 |
| Yogurt | 50 g/day | 0.92 (0.88,0.96) | 0.00036798 |
| Folate | 100 µg/day | 0.88 (0.82,0.95) | 0.00117343 |
| Carbohydrates | 50 g/day | 0.89 (0.83,0.96) | 0.00128295 |
| Total sugars | 50 g/day | 0.88 (0.81,0.95) | 0.00154247 |
| Red and processed meat | 30 g/day | 1.08 (1.03,1.12) | 0.00161454 |
| Fruit | 200 g/day | 0.90 (0.85,0.96) | 0.00191377 |
| Vitamin C | 100 mg/day | 0.90 (0.83,0.96) | 0.00237446 |
| Breakfast cereal | 40 g/day | 0.93 (0.89,0.98) | 0.00359132 |
| Fibre | 5 g/day | 0.92 (0.86,0.97) | 0.00387306 |
| Potassium | 1000 mg/day | 0.89 (0.82,0.97) | 0.00506577 |
| **Not FDR significant** |  |  |  |
| Chocolate | 10 g/day | 1.09 (1.02,1.16) | 0.01131481 |
| White bread | 10 g/day | 1.08 (1.01,1.15) | 0.01566167 |
| Dried fruit | 40 g/day | 0.79 (0.65,0.97) | 0.02151880 |
| Gravy/cream cheese sauce | 10 g/day | 1.10 (1.01,1.20) | 0.02306702 |
| Processed meat | 10 g/day | 1.07 (1.01,1.13) | 0.02309171 |
| Vitamin B6 | 1 mg/day | 0.84 (0.72,0.98) | 0.02671811 |
| Red meat | 20 g/day | 1.05 (1.00,1.09) | 0.03024790 |
| Zinc | 2 mg/day | 0.92 (0.85,0.99) | 0.03383249 |
| Tea | 500 g/day | 0.96 (0.92,1.00) | 0.03439078 |
| Carotene | 1000 µg/day | 0.96 (0.92,1.00) | 0.03960963 |
| Fruit squash | 50 g/day | 1.05 (1.00,1.10) | 0.04060006 |
| Green/red peppers | >1 wk vs <1 wk | 0.96 (0.93,1.00) | 0.05553418 |
| Vegetables | 100 g/day | 0.96 (0.91,1.00) | 0.06919334 |
| Brown bread | 10 g/day | 0.94 (0.87,1.01) | 0.07455062 |
| Boiled sweets | 5 g/day | 0.91 (0.83,1.01) | 0.08275008 |
| Free sugars | 30 g/day | 0.94 (0.88,1.01) | 0.08406961 |
| Eggs | 10 g/day | 1.03 (0.99,1.08) | 0.08856211 |
| Apples | >1 wk vs <1 wk | 0.97 (0.93,1.01) | 0.10913890 |
| Baked beans | >1 wk vs <1 wk | 0.97 (0.94,1.01) | 0.11891682 |
| Oranges | >1 wk vs <1 wk | 0.97 (0.94,1.01) | 0.13471088 |
| Protein | 15 g/day | 0.94 (0.87,1.02) | 0.14168429 |
| Saturated fat | 10 g/day | 1.05 (0.98,1.13) | 0.14767937 |
| Stewed fruit | >1 wk vs <1 wk | 0.97 (0.94,1.01) | 0.19291350 |
| Vitamin E | 2 mg/day | 1.05 (0.97,1.13) | 0.20526452 |
| Pears | >1 wk vs <1 wk | 1.02 (0.99,1.06) | 0.21530932 |
| Fizzy drink | 50 g/day | 1.05 (0.97,1.13) | 0.21706538 |
| Green peas | >1 wk vs <1 wk | 1.02 (0.98,1.06) | 0.25737879 |
| Bananas | >1 wk vs <1 wk | 0.97 (0.93,1.02) | 0.26102513 |
| Polyunsaturated fat | 5 g/day | 1.06 (0.95,1.17) | 0.30466068 |
| Swede | >1 wk vs <1 wk | 0.98 (0.94,1.02) | 0.31197585 |
| Cucumbers | >1 wk vs <1 wk | 0.98 (0.95,1.02) | 0.31450970 |
| Green beans | >1 wk vs <1 wk | 1.02 (0.98,1.06) | 0.32712814 |
| Onion | >1 wk vs <1 wk | 0.98 (0.93,1.02) | 0.33060259 |
| Fruit juice | 100 g/day | 0.98 (0.93,1.03) | 0.35080555 |
| Broccoli | >1 wk vs <1 wk | 0.98 (0.94,1.02) | 0.37144062 |
| Soup | 40 g/day | 0.97 (0.92,1.03) | 0.37509573 |
| Chips | 25 g/day | 1.02 (0.97,1.08) | 0.38687402 |
| Celery | >1 wk vs <1 wk | 0.99 (0.95,1.02) | 0.44361346 |
| Beetroot | >1 wk vs <1 wk | 1.01 (0.98,1.05) | 0.46975507 |
| Courgettes | >1 wk vs <1 wk | 0.99 (0.94,1.03) | 0.49811442 |
| Tomatoes | >1 wk vs <1 wk | 1.02 (0.96,1.09) | 0.51332947 |
| Cabbage | >1 wk vs <1 wk | 1.01 (0.98,1.05) | 0.51416708 |
| Spinach | >1 wk vs <1 wk | 0.98 (0.93,1.04) | 0.52530038 |
| Jam | 5 g/day | 0.97 (0.87,1.08) | 0.52602130 |
| Chick peas/lentils | >1 wk vs <1 wk | 0.98 (0.92,1.04) | 0.53724041 |
| Energy (kj) | 1000 kj/day | 0.99 (0.96,1.02) | 0.54225677 |
| Ice cream | >1 wk vs <1 wk | 0.99 (0.94,1.03) | 0.54887216 |
| Lettuce | >1 wk vs <1 wk | 0.99 (0.95,1.03) | 0.59203027 |
| Carrots | >1 wk vs <1 wk | 1.02 (0.96,1.08) | 0.61366552 |
| Brussels sprouts | >1 wk vs <1 wk | 0.99 (0.95,1.03) | 0.61434528 |
| Mushrooms | >1 wk vs <1 wk | 1.01 (0.97,1.05) | 0.63200818 |
| Sweetcorn | >1 wk vs <1 wk | 0.99 (0.95,1.03) | 0.63264994 |
| Sweet biscuits | 10 g/day | 1.01 (0.95,1.08) | 0.64827347 |
| Soya/tofu | >1 wk vs <1 wk | 1.02 (0.93,1.13) | 0.64849850 |
| Crisps | 10 g/day | 1.02 (0.92,1.14) | 0.65993208 |
| Avocado | >1 wk vs <1 wk | 1.01 (0.95,1.08) | 0.66342691 |
| Cakes and pastries | 10 g/day | 0.99 (0.92,1.05) | 0.66753715 |
| Stone fruit | >1 wk vs <1 wk | 0.99 (0.96,1.03) | 0.67816304 |
| Leeks | >1 wk vs <1 wk | 1.01 (0.97,1.05) | 0.72564717 |
| Crispbread | 10 g/day | 0.99 (0.94,1.04) | 0.75162923 |
| Thiamine | 1 mg/day | 1.00 (0.98, 1.03) | 0.76060528 |
| Vitamin B12 | 2 µg/day | 0.99 (0.94,1.05) | 0.75662649 |
| Non-oily fish | 10 g/day | 0.99 (0.92,1.06) | 0.76508993 |
| Garlic | >1 wk vs <1 wk | 0.99 (0.96,1.03) | 0.78555511 |
| Nuts | 10 g/day | 0.99 (0.92,1.07) | 0.81278348 |
| Pasta | 20 g/day | 0.99 (0.93,1.06) | 0.83399010 |
| Retinol | 250 µg/day | 1.00 (0.95,1.06) | 0.86756575 |
| Parsnip | >1 wk vs <1 wk | 1.00 (0.96,1.04) | 0.88177753 |
| Coffee | 500 g/day | 1.00 (0.95,1.06) | 0.89383674 |
| Grapefruit | >1 wk vs <1 wk | 1.00 (0.96,1.04) | 0.89936258 |
| Soya milk | >1 wk vs <1 wk | 1.01 (0.91,1.11) | 0.90541477 |
| Cheese | 10 g/day | 1.00 (0.96,1.05) | 0.92182738 |
| Oily fish | 10 g/day | 1.00 (0.95,1.05) | 0.92247969 |
| Monounsaturated fat | 5 g/day | 1.00 (0.94,1.07) | 0.92404681 |
| Vitamin D | 1 µg/day | 1.00 (0.95,1.06) | 0.95783436 |
| Total fat | 20 g/day | 1.00 (0.92,1.09) | 0.95875227 |
| Chicken | 20 g/day | 1.00 (0.94,1.07) | 0.97581452 |
| Rice | 10 g/day | 1.00 (0.96,1.04) | 0.98093224 |
| Potatoes | 40 g/day | 1.00 (0.95,1.06) | 0.98856187 |
| Cauliflower | >1 wk vs <1 wk | 1.00 (0.96,1.04) | 0.99984611 |

^1^Associations between each of the 97 foods or nutrients and colorectal cancer incidence separately using Cox proportional hazards regression models stratified by year of birth, date of completion of the 3-year re-survey (which is baseline for this study), and region of residence (10 geographical regions (9 in England), and Scotland), and adjusted for area-based deprivation (fifths, based on the Townsend deprivation score, unknown), highest educational qualification (none, technical, secondary, tertiary, unknown), body mass index (<20, 20-22.49, 22.5-24.9, 25.0-27.49, 27.5-29.9, 30-32.49, 32.5-34.9, 35+ kg/m2, unknown), height (<160, 160–164.9, ≥165 cm, unknown), strenuous exercise (none, ≤ once per week, > once per week, unknown), dietary energy intake (except for the analysis of energy and risk; fifths, unknown), alcohol (none, 1-5, 6-10, ≥ 11 drinks per week, unknown), smoking (never, past, current 1–4, current 5–9, current <10, current 10–14, current 15–19, current 20–24, current 25–29, current ≥30 cigarettes per day, unknown), current use of hormonal therapy for menopause (no, yes, unknown), and family history of bowel cancer (no, yes).

### Supplementary Table 3. Associations of FDR-significant dietary factors with risk of colorectal cancer by level of adjustment for lifestyle factors

| **Food or nutrient** | **Increment** | **Model 1 RR (95% CI)** | **Model 2 RR (95% CI)** | **Fully adjusted RR (95% CI)** |
| --- | --- | --- | --- | --- |
| Alcohol | 20 g/day | 1.16 (1.11,1.21) | 1.15 (1.09,1.20) | 1.15 (1.09,1.20) |
| Calcium | 300 mg/day | 0.88 (0.84,0.93) | 0.88 (0.83,0.93) | 0.83 (0.77,0.89) |
| Dairy milk | 200 g/day | 0.86 (0.81,0.91) | 0.85 (0.80,0.91) | 0.86 (0.81,0.92) |
| Phosphorus | 300 mg/day | 0.92 (0.88,0.97) | 0.92 (0.88,0.97) | 0.84 (0.78,0.91) |
| Riboflavin | 1 mg/day | 0.86 (0.79,0.93) | 0.86 (0.79,0.93) | 0.83 (0.75,0.91) |
| Magnesium | 100 mg/day | 0.91 (0.85,0.97) | 0.90 (0.85,0.97) | 0.84 (0.77,0.92) |
| Wholegrains | 20 g/day | 0.87 (0.83,0.92) | 0.89 (0.85,0.95) | 0.90 (0.85,0.95) |
| Yogurt | 50 g/day | 0.90 (0.86,0.95) | 0.91 (0.87,0.95) | 0.92 (0.88,0.96) |
| Folate | 100 µg/day | 0.90 (0.85,0.96) | 0.91 (0.86,0.97) | 0.88 (0.82,0.95) |
| Carbohydrates | 50 g/day | 0.94 (0.90,0.97) | 0.94 (0.91,0.98) | 0.89 (0.83,0.96) |
| Total sugars | 50 g/day | 0.89 (0.84,0.94) | 0.90 (0.85,0.96) | 0.88 (0.81,0.95) |
| Red/processed meat | 30 g/day | 1.09 (1.04,1.14) | 1.07 (1.03,1.12) | 1.08 (1.03,1.12) |
| Fruit | 200 g/day | 0.86 (0.81,0.92) | 0.89 (0.84,0.95) | 0.90 (0.85,0.96) |
| Vitamin C | 100 mg/day | 0.88 (0.82,0.94) | 0.90 (0.84,0.96) | 0.90 (0.83,0.96) |
| Breakfast cereal | 40 g/day | 0.90 (0.86,0.94) | 0.92 (0.88,0.96) | 0.93 (0.89,0.98) |
| Fibre | 5 g/day | 0.90 (0.86,0.95) | 0.92 (0.88,0.97) | 0.92 (0.86,0.97) |
| Potassium | 1000 mg/day | 0.94 (0.88,1.00) | 0.93 (0.87,0.99) | 0.89 (0.82,0.97) |

Model 1: associations between each of the 17 foods or nutrients and colorectal cancer incidence stratified by year of birth, date of completion of the 3-year re-survey (which is baseline for this study), and region of residence (10 geographical regions (9 in England), and Scotland); Model 2: associations additionally adjusted for area-based deprivation (fifths, based on the Townsend deprivation score, unknown), highest educational qualification (none, technical, secondary, tertiary, unknown), body mass index (<20, 20-22.49, 22.5-24.9, 25.0-27.49, 27.5-29.9, 30-32.49, 32.5-34.9, 35+ kg/m2, unknown), height (<160, 160–164.9, ≥165 cm, unknown), strenuous exercise (none, ≤ once per week, > once per week, unknown), smoking (never, past, current 1–4, current 5–9, current <10, current 10–14, current 15–19, current 20–24, current 25–29, current ≥30 cigarettes per day, unknown), current use of hormonal therapy for menopause (no, yes, unknown), and family history of bowel cancer (no, yes); Fully adjusted RR: associations additionally adjusted for dietary energy intake (except for the analysis of energy and risk; fifths, unknown), alcohol (none, 1-5, 6-10, ≥ 11 drinks per week, unknown).

| Supplementary Table 4. Associations of FDR-significant dietary factors and colorectal cancer risk by self-reported health and follow-up period | | | | | | |
| --- | --- | --- | --- | --- | --- | --- |
|  | **All participants** | | **Participants with self-reported good or excellent health at baseline** | | **Excluding the first 5 years of follow-up** | |
| **Dietary factors, daily intakes unless otherwise specified** | **Cases** | **RR^1^ (95% CI)** | **Cases** | **RR^1^ (95% CI)** | **Cases** | **RR^1^ (95% CI)** |
| **Alcohol, units/wk, g** |  |  |  |  |  |  |
| 0 | 4262 | 1 | 3078 | 1 | 3463 | 1 |
| 1-5 | 3735 | 0.97 (0.93,1.01) | 3041 | 0.98 (0.93,1.03) | 3061 | 0.97 (0.92,1.02) |
| 6-10 | 2501 | 1.04 (0.99,1.09) | 2055 | 1.03 (0.97,1.09) | 2074 | 1.06 (1.00,1.12) |
| 11+ | 1753 | 1.17 (1.10,1.24) | 1490 | 1.18 (1.11,1.26) | 1427 | 1.17 (1.09,1.25) |
| Trend per 20g/day |  | 1.15 (1.09,1.20) |  | 1.15 (1.09,1.21) |  | 1.15 (1.09,1.21) |
| **Calcium, mg** |  |  |  |  |  |  |
| lowest quintile | 2533 | 1 | 1989 | 1 | 2074 | 1 |
| quintile 2 | 2448 | 0.93 (0.87,0.98) | 1915 | 0.89 (0.84,0.96) | 1989 | 0.92 (0.86,0.98) |
| quintile 3 | 2489 | 0.91 (0.86,0.97) | 1959 | 0.88 (0.82,0.94) | 2015 | 0.90 (0.84,0.96) |
| quintile 4 | 2387 | 0.85 (0.80,0.91) | 1907 | 0.83 (0.77,0.89) | 1989 | 0.87 (0.80,0.93) |
| highest quintile | 2394 | 0.83 (0.77,0.90) | 1894 | 0.81 (0.74,0.88) | 1958 | 0.83 (0.77,0.90) |
| Trends per 300 mg/day |  | 0.83 (0.77,0.89) |  | 0.81 (0.75,0.88) |  | 0.84 (0.78,0.90) |
| **Dairy milk, g** |  |  |  |  |  |  |
| lowest quintile | 2559 | 1 | 2071 | 1 | 2087 | 1 |
| quintile 2 | 2559 | 0.97 (0.91,1.02) | 2062 | 0.97 (0.91,1.03) | 2077 | 0.96 (0.90,1.02) |
| quintile 3 | 2372 | 0.92 (0.87,0.98) | 1855 | 0.91 (0.85,0.97) | 1951 | 0.93 (0.87,0.99) |
| quintile 4 | 2476 | 0.90 (0.85,0.95) | 1927 | 0.88 (0.83,0.94) | 2033 | 0.91 (0.85,0.96) |
| highest quintile | 2285 | 0.89 (0.84,0.94) | 1749 | 0.88 (0.82,0.94) | 1877 | 0.89 (0.84,0.95) |
| Trend per 200g/day |  | 0.86 (0.81,0.92) |  | 0.85 (0.79,0.91) |  | 0.87 (0.81,0.93) |
| **Phosphorus, mg** |  |  |  |  |  |  |
| lowest quintile | 2495 | 1 | 1903 | 1 | 2052 | 1 |
| quintile 2 | 2478 | 0.94 (0.88,1.00) | 1938 | 0.92 (0.86,0.99) | 1994 | 0.92 (0.85,0.98) |
| quintile 3 | 2394 | 0.88 (0.82,0.94) | 1918 | 0.86 (0.79,0.93) | 1983 | 0.88 (0.81,0.95) |
| quintile 4 | 2492 | 0.88 (0.82,0.95) | 1985 | 0.85 (0.78,0.93) | 2032 | 0.87 (0.80,0.95) |
| highest quintile | 2392 | 0.82 (0.75,0.90) | 1920 | 0.79 (0.72,0.87) | 1964 | 0.82 (0.74,0.90) |
| Trend per 300 mg/day |  | 0.84 (0.78,0.91) |  | 0.82 (0.75,0.89) |  | 0.84 (0.78,0.92) |
| **Magnesium, mg** |  |  |  |  |  |  |
| lowest quintile | 2490 | 1 | 1850 | 1 | 2028 | 1 |
| quintile 2 | 2476 | 0.95 (0.90,1.01) | 1960 | 0.95 (0.89,1.02) | 2036 | 0.96 (0.90,1.02) |
| quintile 3 | 2477 | 0.93 (0.88,0.99) | 1951 | 0.90 (0.84,0.97) | 2039 | 0.94 (0.87,1.01) |
| quintile 4 | 2411 | 0.89 (0.83,0.96) | 1967 | 0.88 (0.81,0.95) | 1961 | 0.88 (0.82,0.95) |
| highest quintile | 2397 | 0.87 (0.81,0.94) | 1936 | 0.84 (0.78,0.92) | 1961 | 0.86 (0.79,0.94) |
| Trend per 100 mg/day |  | 0.84 (0.77,0.92) |  | 0.81 (0.73,0.90) |  | 0.83 (0.75,0.91) |
| **Wholegrains, g** |  |  |  |  |  |  |
| lowest quintile | 2540 | 1 | 1904 | 1 | 2117 | 1 |
| quintile 2 | 2451 | 0.95 (0.90,1.01) | 1917 | 0.95 (0.89,1.01) | 1989 | 0.93 (0.87,0.99) |
| quintile 3 | 2504 | 0.97 (0.91,1.02) | 1979 | 0.95 (0.89,1.01) | 2034 | 0.94 (0.89,1.00) |
| quintile 4 | 2329 | 0.89 (0.84,0.94) | 1890 | 0.88 (0.83,0.94) | 1899 | 0.87 (0.82,0.93) |
| highest quintile | 2427 | 0.91 (0.86,0.96) | 1974 | 0.90 (0.84,0.96) | 1986 | 0.89 (0.83,0.95) |
| Trend per 20g/day |  | 0.90 (0.85,0.95) |  | 0.89 (0.83,0.95) |  | 0.88 (0.83,0.94) |
| **Yogurt, number pw** |  |  |  |  |  |  |
| <=2 | 5519 | 1 | 4275 | 1 | 4531 | 1 |
| 3-4 | 2355 | 0.94 (0.89,0.98) | 1845 | 0.92 (0.87,0.98) | 1928 | 0.93 (0.88,0.98) |
| 5-6 | 1816 | 0.94 (0.89,0.99) | 1472 | 0.94 (0.89,1.00) | 1490 | 0.94 (0.88,1.00) |
| 7+ | 2561 | 0.92 (0.88,0.97) | 2072 | 0.92 (0.88,0.98) | 2076 | 0.91 (0.86,0.96) |
| Trend per 50g/day |  | 0.92 (0.88,0.96) |  | 0.93 (0.88,0.97) |  | 0.91 (0.87,0.96) |
| **Folate, µg** |  |  |  |  |  |  |
| lowest quintile | 2460 | 1 | 1859 | 1 | 2003 | 1 |
| quintile 2 | 2411 | 0.94 (0.89,1.00) | 1898 | 0.94 (0.88,1.00) | 1989 | 0.95 (0.89,1.02) |
| quintile 3 | 2497 | 0.96 (0.90,1.02) | 1979 | 0.95 (0.88,1.01) | 2044 | 0.96 (0.90,1.03) |
| quintile 4 | 2463 | 0.92 (0.87,0.99) | 1974 | 0.92 (0.85,0.99) | 2009 | 0.93 (0.86,0.99) |
| highest quintile | 2420 | 0.89 (0.83,0.95) | 1954 | 0.88 (0.82,0.95) | 1980 | 0.89 (0.83,0.96) |
| Trend per 100 µg/day |  | 0.88 (0.82,0.95) |  | 0.88 (0.81,0.96) |  | 0.88 (0.81,0.96) |
| **Carbohydrates, g** |  |  |  |  |  |  |
| lowest quintile | 2517 | 1 | 1962 | 1 | 2071 | 1 |
| quintile 2 | 2417 | 0.91 (0.85,0.97) | 1933 | 0.92 (0.86,0.99) | 1976 | 0.89 (0.83,0.95) |
| quintile 3 | 2434 | 0.88 (0.82,0.95) | 1953 | 0.91 (0.83,0.98) | 1980 | 0.86 (0.79,0.93) |
| quintile 4 | 2392 | 0.84 (0.78,0.92) | 1881 | 0.86 (0.79,0.95) | 1938 | 0.82 (0.75,0.90) |
| highest quintile | 2491 | 0.86 (0.78,0.95) | 1935 | 0.90 (0.81,1.01) | 2060 | 0.85 (0.77,0.95) |
| Trend per 50g/day |  | 0.89 (0.83,0.96) |  | 0.92 (0.85,1.00) |  | 0.88 (0.82,0.95) |
| **Total sugars, g** |  |  |  |  |  |  |
| lowest quintile | 2509 | 1 | 1942 | 1 | 2050 | 1 |
| quintile 2 | 2491 | 0.96 (0.91,1.02) | 1990 | 0.97 (0.91,1.03) | 2067 | 0.97 (0.91,1.04) |
| quintile 3 | 2399 | 0.91 (0.86,0.97) | 1924 | 0.92 (0.85,0.98) | 1930 | 0.90 (0.84,0.96) |
| quintile 4 | 2405 | 0.90 (0.84,0.96) | 1912 | 0.90 (0.84,0.97) | 1961 | 0.89 (0.83,0.96) |
| highest quintile | 2447 | 0.90 (0.84,0.97) | 1896 | 0.91 (0.84,0.99) | 2017 | 0.90 (0.83,0.98) |
| Trend per 50 g/day |  | 0.88 (0.81,0.95) |  | 0.89 (0.81,0.97) |  | 0.87 (0.80,0.95) |
| **Red/processed meat, g** |  |  |  |  |  |  |
| lowest quintile | 2268 | 1 | 1787 | 1 | 1872 | 1 |
| quintile 2 | 2498 | 1.08 (1.02,1.15) | 1974 | 1.10 (1.03,1.17) | 2033 | 1.07 (1.00,1.14) |
| quintile 3 | 2470 | 1.07 (1.00,1.13) | 1932 | 1.06 (0.99,1.13) | 1991 | 1.04 (0.98,1.11) |
| quintile 4 | 2542 | 1.10 (1.04,1.17) | 2016 | 1.10 (1.03,1.18) | 2086 | 1.09 (1.02,1.16) |
| highest quintile | 2473 | 1.09 (1.03,1.16) | 1955 | 1.08 (1.00,1.15) | 2043 | 1.09 (1.02,1.16) |
| Trend per 30 g/day |  | 1.08 (1.03,1.12) |  | 1.06 (1.01,1.12) |  | 1.07 (1.02,1.12) |
| **Fruit pw** |  |  |  |  |  |  |
| <=5 | 2069 | 1 | 1486 | 1 | 1682 | 1 |
| 6-9 | 2356 | 0.98 (0.92,1.04) | 1780 | 0.97 (0.91,1.04) | 1937 | 0.99 (0.93,1.06) |
| 10-14 | 2992 | 0.95 (0.90,1.01) | 2410 | 0.97 (0.91,1.03) | 2449 | 0.96 (0.90,1.02) |
| 15-19 | 1570 | 0.92 (0.86,0.98) | 1285 | 0.92 (0.85,0.99) | 1272 | 0.92 (0.85,0.99) |
| 20+ | 3264 | 0.92 (0.87,0.98) | 2703 | 0.92 (0.86,0.99) | 2685 | 0.93 (0.87,1.00) |
| Trend per 200g/day |  | 0.90 (0.85,0.96) |  | 0.91 (0.85,0.98) |  | 0.91 (0.85,0.98) |
| **Vitamin C, mg** |  |  |  |  |  |  |
| lowest quintile | 2578 | 1 | 1877 | 1 | 2109 | 1 |
| quintile 2 | 2434 | 0.93 (0.88,0.99) | 1856 | 0.92 (0.86,0.98) | 1976 | 0.92 (0.87,0.98) |
| quintile 3 | 2432 | 0.92 (0.87,0.98) | 1981 | 0.94 (0.88,1.00) | 2012 | 0.93 (0.88,0.99) |
| quintile 4 | 2461 | 0.93 (0.88,0.99) | 2018 | 0.94 (0.88,1.00) | 2004 | 0.93 (0.87,0.99) |
| highest quintile | 2346 | 0.90 (0.85,0.96) | 1932 | 0.90 (0.84,0.96) | 1924 | 0.90 (0.84,0.96) |
| Trend per 100 mg/day |  | 0.90 (0.83,0.96) |  | 0.90 (0.83,0.98) |  | 0.90 (0.83,0.97) |
| **Riboflavin, mg** |  |  |  |  |  |  |
| lowest quintile | 2540 | 1 | 1988 | 1 | 2082 | 1 |
| quintile 2 | 2486 | 0.99 (0.94,1.05) | 1955 | 0.97 (0.91,1.03) | 2023 | 0.98 (0.92,1.05) |
| quintile 3 | 2472 | 0.95 (0.89,1.01) | 1932 | 0.91 (0.85,0.97) | 2023 | 0.95 (0.89,1.01) |
| quintile 4 | 2367 | 0.92 (0.86,0.98) | 1900 | 0.90 (0.84,0.96) | 1922 | 0.91 (0.85,0.98) |
| highest quintile | 2386 | 0.89 (0.83,0.95) | 1889 | 0.86 (0.80,0.93) | 1975 | 0.90 (0.83,0.97) |
| Trend per 1mg/day |  | 0.83 (0.75,0.91) |  | 0.79 (0.71,0.88) |  | 0.84 (0.75,0.93) |
| **Cereal, bowls pw, g** |  |  |  |  |  |  |
| <7 | 6964 | 1 | 5429 | 1 | 5735 | 1 |
| 7 | 5139 | 0.95 (0.91,0.98) | 4124 | 0.96 (0.92,1.00) | 4170 | 0.94 (0.90,0.98) |
| 8+ | 148 | 0.92 (0.78,1.08) | 111 | 0.92 (0.76,1.12) | 120 | 0.91 (0.76,1.09) |
| Trend per 40 g/day |  | 0.93 (0.89,0.98) |  | 0.95 (0.90,1.00) |  | 0.92 (0.87,0.97) |
| **Fibre, g per day** |  |  |  |  |  |  |
| lowest quintile | 2456 | 1 | 1818 | 1 | 2040 | 1 |
| quintile 2 | 2524 | 0.99 (0.94,1.05) | 1944 | 0.98 (0.91,1.04) | 2043 | 0.96 (0.90,1.03) |
| quintile 3 | 2429 | 0.95 (0.89,1.01) | 1966 | 0.95 (0.89,1.02) | 1986 | 0.92 (0.86,0.99) |
| quintile 4 | 2453 | 0.95 (0.89,1.01) | 1966 | 0.93 (0.86,1.00) | 2004 | 0.92 (0.86,0.99) |
| highest quintile | 2389 | 0.91 (0.85,0.98) | 1970 | 0.91 (0.84,0.98) | 1952 | 0.89 (0.83,0.96) |
| Trend per 5 g/day |  | 0.92 (0.86,0.97) |  | 0.91 (0.86,0.98) |  | 0.90 (0.84,0.96) |
| **Potassium, mg** |  |  |  |  |  |  |
| lowest quintile | 2462 | 1 | 1844 | 1 | 2006 | 1 |
| quintile 2 | 2459 | 0.95 (0.90,1.01) | 1925 | 0.95 (0.88,1.01) | 2026 | 0.96 (0.90,1.03) |
| quintile 3 | 2431 | 0.93 (0.87,0.99) | 1937 | 0.91 (0.85,0.98) | 1967 | 0.91 (0.85,0.98) |
| quintile 4 | 2468 | 0.92 (0.86,0.99) | 1994 | 0.91 (0.84,0.98) | 2018 | 0.92 (0.85,0.99) |
| highest quintile | 2431 | 0.90 (0.83,0.97) | 1964 | 0.88 (0.81,0.95) | 2008 | 0.90 (0.83,0.98) |
| Trend 1000 mg/day |  | 0.89 (0.82,0.97) |  | 0.87 (0.79,0.95) |  | 0.89 (0.82,0.98) |

**^1^**Associations between each of the 17 foods or nutrients and colorectal cancer incidence separately using Cox proportional hazards regression models stratified by year of birth, date of completion of the 3-year re-survey (which is baseline for this study), and region of residence (10 geographical regions (9 in England), and Scotland), and adjusted for area-based deprivation (fifths, based on the Townsend deprivation score, unknown), highest educational qualification (none, technical, secondary, tertiary, unknown), body mass index (<20, 20-22.49, 22.5-24.9, 25.0-27.49, 27.5-29.9, 30-32.49, 32.5-34.9, 35+ kg/m2, unknown), height (<160, 160–164.9, ≥165 cm, unknown), strenuous exercise (none, ≤ once per week, > once per week, unknown), dietary energy intake (except for the analysis of energy and risk; fifths, unknown), alcohol (none, 1-5, 6-10, ≥ 11 drinks per week, unknown), smoking (never, past, current 1–4, current 5–9, current <10, current 10–14, current 15–19, current 20–24, current 25–29, current ≥30 cigarettes per day, unknown), current use of hormonal therapy for menopause (no, yes, unknown), and family history of bowel cancer (no, yes).

| Supplementary Table 5. Associations of FDR-significant dietary factors and colorectal cancer risk by cancer site | | | | | | |  |
| --- | --- | --- | --- | --- | --- | --- | --- |
|  | **Proximal** | | **Distal** | | **Rectum** | |  |
| **Dietary factors, daily intakes unless otherwise specified** | **Cases** | **RR^1^ (95% CI)** | **Cases** | **RR^1^ (95% CI)** | **Cases** | **RR^1^ (95% CI)** | **Phet** |
| **Alcohol, units/wk, g** |  |  |  |  |  |  |  |
| 0 | 1866 | 1 | 1090 | 1 | 1125 | 1 |  |
| 1-5 | 1666 | 0.99 (0.93,1.06) | 941 | 0.94 (0.86,1.03) | 982 | 0.96 (0.88,1.04) |  |
| 6-10 | 979 | 0.94 (0.87,1.02) | 682 | 1.09 (0.99,1.21) | 732 | 1.13 (1.03,1.25) |  |
| 11+ | 713 | 1.11 (1.02,1.22) | 451 | 1.16 (1.04,1.30) | 513 | 1.26 (1.13,1.41) |  |
| Trend per 20g/day |  | 1.06 (0.99,1.14) |  | 1.17 (1.06,1.28) |  | 1.25 (1.15,1.37) | 0.02 |
| **Calcium, mg** |  |  |  |  |  |  |  |
| lowest quintile | 1054 | 1 | 672 | 1 | 705 | 1 |  |
| quintile 2 | 1024 | 0.92 (0.84,1.00) | 620 | 0.88 (0.79,0.99) | 715 | 1.00 (0.89,1.11) |  |
| quintile 3 | 1066 | 0.91 (0.83,1.00) | 648 | 0.90 (0.80,1.02) | 657 | 0.90 (0.80,1.01) |  |
| quintile 4 | 1058 | 0.88 (0.79,0.97) | 599 | 0.82 (0.72,0.93) | 641 | 0.86 (0.76,0.97) |  |
| highest quintile | 1022 | 0.82 (0.73,0.91) | 625 | 0.86 (0.75,1.00) | 634 | 0.82 (0.71,0.94) |  |
| Trends per 300 mg/day |  | 0.83 (0.74,0.92) |  | 0.86 (0.75,0.99) |  | 0.80 (0.70,0.92) | 0.77 |
| **Dairy milk, g** |  |  |  |  |  |  |  |
| lowest quintile | 1083 | 1 | 669 | 1 | 713 | 1 |  |
| quintile 2 | 1039 | 0.92 (0.85,1.00) | 658 | 0.95 (0.85,1.06) | 743 | 1.02 (0.92,1.13) |  |
| quintile 3 | 1039 | 0.95 (0.87,1.03) | 609 | 0.91 (0.81,1.02) | 644 | 0.92 (0.82,1.02) |  |
| quintile 4 | 1083 | 0.92 (0.84,1.00) | 648 | 0.90 (0.81,1.01) | 643 | 0.87 (0.78,0.97) |  |
| highest quintile | 980 | 0.88 (0.80,0.96) | 580 | 0.87 (0.78,0.98) | 609 | 0.88 (0.79,0.99) |  |
| Trend per 200g/day |  | 0.87 (0.79,0.96) |  | 0.85 (0.75,0.97) |  | 0.83 (0.73,0.94) | 0.85 |
| **Phosphorus, mg** |  |  |  |  |  |  |  |
| lowest quintile | 1034 | 1 | 650 | 1 | 713 | 1 |  |
| quintile 2 | 1063 | 0.96 (0.87,1.06) | 662 | 0.95 (0.84,1.07) | 671 | 0.92 (0.81,1.03) |  |
| quintile 3 | 986 | 0.85 (0.76,0.95) | 632 | 0.88 (0.76,1.00) | 665 | 0.89 (0.78,1.01) |  |
| quintile 4 | 1103 | 0.92 (0.82,1.04) | 616 | 0.83 (0.71,0.96) | 657 | 0.85 (0.74,0.99) |  |
| highest quintile | 1038 | 0.84 (0.74,0.96) | 604 | 0.81 (0.68,0.95) | 646 | 0.79 (0.67,0.94) |  |
| Trend per 300 mg/day |  | 0.87 (0.77,0.97) |  | 0.81 (0.70,0.94) |  | 0.82 (0.71,0.95) | 0.75 |
| **Magnesium, mg** |  |  |  |  |  |  |  |
| lowest quintile | 1039 | 1 | 648 | 1 | 707 | 1 |  |
| quintile 2 | 1021 | 0.94 (0.85,1.03) | 684 | 1.00 (0.89,1.13) | 676 | 0.93 (0.83,1.04) |  |
| quintile 3 | 1070 | 0.96 (0.87,1.06) | 643 | 0.92 (0.81,1.04) | 663 | 0.90 (0.79,1.01) |  |
| quintile 4 | 1081 | 0.95 (0.86,1.06) | 582 | 0.81 (0.71,0.93) | 641 | 0.85 (0.75,0.97) |  |
| highest quintile | 1013 | 0.88 (0.78,0.99) | 607 | 0.85 (0.73,0.98) | 665 | 0.85 (0.74,0.99) |  |
| Trend per 100 mg/day |  | 0.88 (0.77,1.01) |  | 0.76 (0.64,0.91) |  | 0.82 (0.69,0.98) | 0.48 |
| **Wholegrains, g** |  |  |  |  |  |  |  |
| lowest quintile | 1046 | 1 | 692 | 1 | 697 | 1 |  |
| quintile 2 | 1054 | 0.99 (0.91,1.08) | 608 | 0.87 (0.78,0.97) | 688 | 0.98 (0.88,1.09) |  |
| quintile 3 | 1048 | 0.97 (0.89,1.06) | 675 | 0.96 (0.86,1.07) | 671 | 0.95 (0.85,1.06) |  |
| quintile 4 | 1035 | 0.95 (0.87,1.04) | 583 | 0.82 (0.73,0.92) | 635 | 0.90 (0.80,1.00) |  |
| highest quintile | 1041 | 0.93 (0.85,1.02) | 606 | 0.84 (0.75,0.95) | 661 | 0.92 (0.82,1.03) |  |
| Trend per 20g/day |  | 0.92 (0.84,1.01) |  | 0.84 (0.75,0.94) |  | 0.90 (0.80,1.00) | 0.48 |
| **Yogurt, number pw, g** |  |  |  |  |  |  |  |
| <=2 | 2336 | 1 | 1432 | 1 | 1521 | 1 |  |
| 3-4 | 1000 | 0.93 (0.87,1.01) | 596 | 0.91 (0.83,1.01) | 659 | 0.96 (0.87,1.05) |  |
| 5-6 | 773 | 0.94 (0.87,1.02) | 477 | 0.95 (0.85,1.05) | 485 | 0.92 (0.83,1.02) |  |
| 7+ | 1115 | 0.93 (0.86,1.00) | 659 | 0.92 (0.84,1.01) | 687 | 0.91 (0.83,1.00) |  |
| Trend per 50g/day |  | 0.93 (0.87,1.00) |  | 0.92 (0.84,1.01) |  | 0.91 (0.83,0.99) | 0.94 |
| **Folate, µg** |  |  |  |  |  |  |  |
| lowest quintile | 1013 | 1 | 666 | 1 | 673 | 1 |  |
| quintile 2 | 988 | 0.93 (0.85,1.02) | 639 | 0.91 (0.81,1.02) | 689 | 1.02 (0.91,1.14) |  |
| quintile 3 | 1102 | 1.01 (0.92,1.11) | 636 | 0.89 (0.79,1.00) | 660 | 0.97 (0.86,1.09) |  |
| quintile 4 | 1075 | 0.96 (0.87,1.06) | 633 | 0.86 (0.76,0.98) | 660 | 0.96 (0.85,1.09) |  |
| highest quintile | 1046 | 0.92 (0.82,1.02) | 590 | 0.79 (0.69,0.91) | 670 | 0.96 (0.84,1.09) |  |
| Trend per 100 µg |  | 0.93 (0.82,1.04) |  | 0.78 (0.67,0.90) |  | 0.93 (0.81,1.08) | 0.14 |
| **Carbohydrates, g** |  |  |  |  |  |  |  |
| lowest quintile | 1044 | 1 | 657 | 1 | 713 | 1 |  |
| quintile 2 | 1031 | 0.91 (0.83,1.01) | 635 | 0.90 (0.80,1.02) | 673 | 0.93 (0.83,1.05) |  |
| quintile 3 | 1020 | 0.86 (0.77,0.96) | 654 | 0.89 (0.77,1.03) | 655 | 0.91 (0.79,1.05) |  |
| quintile 4 | 1046 | 0.85 (0.75,0.96) | 607 | 0.80 (0.68,0.94) | 638 | 0.87 (0.75,1.02) |  |
| highest quintile | 1083 | 0.86 (0.75,1.00) | 611 | 0.80 (0.66,0.97) | 673 | 0.89 (0.74,1.06) |  |
| Trend per 50g/day |  | 0.89 (0.80,0.99) |  | 0.85 (0.74,0.97) |  | 0.91 (0.80,1.04) | 0.73 |
| **Total sugars, g** |  |  |  |  |  |  |  |
| lowest quintile | 1052 | 1 | 663 | 1 | 696 | 1 |  |
| quintile 2 | 1051 | 0.95 (0.87,1.04) | 668 | 0.97 (0.86,1.08) | 680 | 0.98 (0.88,1.10) |  |
| quintile 3 | 1000 | 0.88 (0.80,0.97) | 605 | 0.87 (0.77,0.98) | 696 | 1.01 (0.90,1.14) |  |
| quintile 4 | 1060 | 0.91 (0.82,1.00) | 618 | 0.88 (0.77,1.00) | 616 | 0.89 (0.78,1.01) |  |
| highest quintile | 1061 | 0.89 (0.79,0.99) | 610 | 0.87 (0.75,1.00) | 664 | 0.94 (0.82,1.08) |  |
| Trend per 50g/day |  | 0.87 (0.77,0.99) |  | 0.84 (0.72,0.98) |  | 0.91 (0.78,1.06) | 0.77 |
| **Red/processed meat,g** |  |  |  |  |  |  |  |
| lowest quintile | 960 | 1 | 597 | 1 | 619 | 1 |  |
| quintile 2 | 1052 | 1.07 (0.98,1.16) | 641 | 1.06 (0.95,1.18) | 697 | 1.13 (1.02,1.26) |  |
| quintile 3 | 1023 | 1.03 (0.94,1.12) | 676 | 1.11 (0.99,1.24) | 656 | 1.06 (0.95,1.19) |  |
| quintile 4 | 1127 | 1.14 (1.04,1.25) | 611 | 1.01 (0.89,1.13) | 699 | 1.14 (1.02,1.27) |  |
| highest quintile | 1062 | 1.10 (1.00,1.21) | 639 | 1.07 (0.95,1.21) | 681 | 1.12 (1.00,1.26) |  |
| Trend per 30 g/day |  | 1.09 (1.02,1.17) |  | 1.04 (0.95,1.14) |  | 1.09 (1.00,1.19) | 0.65 |
| **Fruit pw** |  |  |  |  |  |  |  |
| <=5 | 855 | 1 | 551 | 1 | 573 | 1 |  |
| 6-9 | 1017 | 1.01 (0.92,1.11) | 604 | 0.95 (0.84,1.06) | 628 | 0.96 (0.86,1.08) |  |
| 10-14 | 1283 | 0.97 (0.89,1.06) | 777 | 0.93 (0.83,1.05) | 810 | 0.95 (0.85,1.06) |  |
| 15-19 | 669 | 0.92 (0.83,1.03) | 399 | 0.88 (0.77,1.01) | 445 | 0.96 (0.84,1.09) |  |
| 20+ | 1400 | 0.93 (0.85,1.02) | 833 | 0.90 (0.80,1.01) | 896 | 0.93 (0.83,1.04) |  |
| Trend per 200 g/day |  | 0.90 (0.82,0.99) |  | 0.89 (0.78,1.01) |  | 0.94 (0.83,1.06) | 0.84 |
| **Vitamin C, mg** |  |  |  |  |  |  |  |
| lowest quintile | 1086 | 1 | 680 | 1 | 704 | 1 |  |
| quintile 2 | 1025 | 0.93 (0.85,1.01) | 638 | 0.92 (0.83,1.03) | 679 | 0.96 (0.86,1.07) |  |
| quintile 3 | 1052 | 0.94 (0.86,1.02) | 656 | 0.94 (0.84,1.05) | 607 | 0.86 (0.76,0.96) |  |
| quintile 4 | 1046 | 0.93 (0.85,1.02) | 617 | 0.89 (0.79,1.00) | 701 | 0.99 (0.89,1.11) |  |
| highest quintile | 1015 | 0.91 (0.83,1.00) | 573 | 0.84 (0.75,0.95) | 661 | 0.94 (0.84,1.05) |  |
| Trend per 100 mg/day |  | 0.91 (0.82,1.02) |  | 0.81 (0.71,0.94) |  | 0.95 (0.83,1.09) | 0.22 |
| **Riboflavin, mg** |  |  |  |  |  |  |  |
| lowest quintile | 1062 | 1 | 653 | 1 | 716 | 1 |  |
| quintile 2 | 1038 | 0.98 (0.89,1.07) | 643 | 1.00 (0.90,1.12) | 714 | 1.03 (0.93,1.15) |  |
| quintile 3 | 1078 | 0.97 (0.88,1.06) | 630 | 0.95 (0.85,1.07) | 664 | 0.93 (0.83,1.05) |  |
| quintile 4 | 1028 | 0.93 (0.84,1.02) | 636 | 0.99 (0.87,1.11) | 609 | 0.87 (0.77,0.98) |  |
| highest quintile | 1018 | 0.88 (0.79,0.97) | 602 | 0.91 (0.80,1.04) | 649 | 0.89 (0.78,1.01) |  |
| Trend per 1mg/day |  | 0.82 (0.71,0.95) |  | 0.88 (0.73,1.07) |  | 0.79 (0.65,0.95) | 0.69 |
| **Cereal, bowls pw** |  |  |  |  |  |  |  |
| <7 | 2934 | 1 | 1813 | 1 | 1933 | 1 |  |
| 7 | 2233 | 0.96 (0.90,1.01) | 1308 | 0.94 (0.87,1.01) | 1382 | 0.94 (0.88,1.01) |  |
| 8+ | 57 | 0.83 (0.64,1.08) | 43 | 1.05 (0.77,1.42) | 37 | 0.83 (0.60,1.15) |  |
| Trend per 40 g/day |  | 0.94 (0.87,1.01) |  | 0.92 (0.84,1.02) |  | 0.92 (0.84,1.01) | 0.93 |
| **Fibre, g** |  |  |  |  |  |  |  |
| lowest quintile | 1008 | 1 | 655 | 1 | 693 | 1 |  |
| quintile 2 | 1067 | 1.01 (0.92,1.11) | 667 | 0.98 (0.88,1.10) | 677 | 0.97 (0.87,1.09) |  |
| quintile 3 | 1054 | 0.98 (0.89,1.08) | 647 | 0.94 (0.84,1.06) | 636 | 0.91 (0.81,1.03) |  |
| quintile 4 | 1078 | 0.99 (0.90,1.09) | 607 | 0.88 (0.77,0.99) | 671 | 0.96 (0.85,1.08) |  |
| highest quintile | 1017 | 0.92 (0.83,1.02) | 588 | 0.85 (0.74,0.97) | 675 | 0.96 (0.85,1.10) |  |
| Trend per 5 g/day |  | 0.93 (0.84,1.01) |  | 0.84 (0.75,0.95) |  | 0.97 (0.87,1.09) | 0.22 |
| **Potassium, mg** |  |  |  |  |  |  |  |
| lowest quintile | 1014 | 1 | 647 | 1 | 701 | 1 |  |
| quintile 2 | 1050 | 0.98 (0.89,1.07) | 653 | 0.96 (0.85,1.08) | 669 | 0.94 (0.84,1.05) |  |
| quintile 3 | 1044 | 0.95 (0.86,1.05) | 636 | 0.91 (0.81,1.04) | 634 | 0.88 (0.78,1.00) |  |
| quintile 4 | 1094 | 0.97 (0.87,1.07) | 625 | 0.88 (0.77,1.01) | 658 | 0.91 (0.79,1.03) |  |
| highest quintile | 1022 | 0.89 (0.79,1.00) | 603 | 0.85 (0.74,0.99) | 690 | 0.93 (0.81,1.08) |  |
| Trend 1000 mg/day |  | 0.88 (0.78,1.00) |  | 0.83 (0.71,0.98) |  | 0.94 (0.80,1.10) | 0.59 |

**^1^**Associations between each of the 17 foods or nutrients and colorectal cancer incidence separately using Cox proportional hazards regression models stratified by year of birth, date of completion of the 3-year re-survey (which is baseline for this study), and region of residence (10 geographical regions (9 in England), and Scotland), and adjusted for area-based deprivation (fifths, based on the Townsend deprivation score, unknown), highest educational qualification (none, technical, secondary, tertiary, unknown), body mass index (<20, 20-22.49, 22.5-24.9, 25.0-27.49, 27.5-29.9, 30-32.49, 32.5-34.9, 35+ kg/m2, unknown), height (<160, 160–164.9, ≥165 cm, unknown), strenuous exercise (none, ≤ once per week, > once per week, unknown), dietary energy intake (except for the analysis of energy and risk; fifths, unknown), alcohol (none, 1-5, 6-10, ≥ 11 drinks per week, unknown), smoking (never, past, current <10, current 10–14, current 15–19, current 20–24, current 25–29, current ≥30 cigarettes per day, unknown), current use of hormonal therapy for menopause (no, yes, unknown), and family history of bowel cancer (no, yes)

### Supplementary Table 6. Relative risks^1^ and 95% confidence intervals for the association of the FDR-significant dietary factors and risk of colorectal cancer stratified by lifestyle factors (excluding alcohol)

|  |  | **Smoking** | |  | **BMI (kg/m^2^)** | |  | **Level of deprivation** | |  | **Alcohol intake** | |  |
| --- | --- | --- | --- | --- | --- | --- | --- | --- | --- | --- | --- | --- | --- |
| Dietary factor | Per daily | Never  N=297,177 | Ever  N=238,033 | Phet | <25  N=241,497 | 25+  N=264.632 | Phet | Highest  N=126,237 | Remainder  N=412,494 | Phet | <7 drinks/wk  N=388,280 | ≥7 drinks/wk  N=154,498 | Phet |
| Alcohol | 20 g | 1.09 (1.01,1.17) | 1.19 (1.12,1.27) | 0.08 | 1.15 (1.07,1.23) | 1.16 (1.08,1.24) | 0.86 | 1.10 (1.00,1.22) | 1.16 (1.10,1.22) | 0.35 | - | - | - |
| Calcium | 300 mg | 0.77 (0.70,0.85) | 0.88 (0.80,0.98) | 0.06 | 0.85 (0.76,0.94) | 0.81 (0.73,0.90) | 0.52 | 0.89 (0.77,1.03) | 0.82 (0.76,0.89) | 0.33 | 0.85 (0.78,0.92) | 0.79 (0.70,0.90) | 0.34 |
| Dairy milk | 200 g | 0.79 (0.72,0.86) | 0.95 (0.86,1.04) | 0.005 | 0.87 (0.79,0.96) | 0.85 (0.78,0.94) | 0.73 | 0.78 (0.68,0.90) | 0.89 (0.83,0.96) | 0.10 | 0.88 (0.82,0.96) | 0.82 (0.73,0.92) | 0.32 |
| Phosphorus | 300 mg | 0.78 (0.70,0.87) | 0.90 (0.81,1.01) | 0.07 | 0.80 (0.71,0.89) | 0.86 (0.77,0.95) | 0.36 | 0.85 (0.72,1.00) | 0.85 (0.78,0.93) | 1.0 | 0.86 (0.79,0.95) | 0.80 (0.69,0.92) | 0.41 |
| Riboflavin | 1 mg | 0.72 (0.63,0.83) | 0.95 (0.83,1.10) | 0.006 | 0.83 (0.72,0.97) | 0.84 (0.73,0.96) | 0.91 | 0.85 (0.69,1.04) | 0.83 (0.75,0.93) | 0.84 | 0.83 (0.74,0.93) | 0.84 (0.70,1.00) | 0.91 |
| Magnesium | 100 mg | 0.78 (0.69,0.88) | 0.91 (0.80,1.04) | 0.09 | 0.76 (0.67,0.88) | 0.86 (0.76,0.97) | 0.19 | 0.82 (0.68,0.99) | 0.85 (0.77,0.94) | 0.74 | 0.86 (0.78,0.96) | 0.80 (0.67,0.95) | 0.49 |
| Wholegrains | 20 g | 0.87 (0.80,0.94) | 0.92 (0.84,1.00) | 0.36 | 0.83 (0.76,0.91) | 0.94 (0.87,1.02) | 0.04 | 0.85 (0.75,0.96) | 0.91 (0.85,0.97) | 0.34 | 0.91 (0.85,0.97) | 0.88 (0.79,0.98) | 0.60 |
| Yogurt | 50 g | 0.88 (0.83,0.94) | 0.96 (0.90,1.03) | 0.06 | 0.91 (0.85,0.98) | 0.92 (0.86,0.98) | 0.82 | 0.96 (0.87,1.05) | 0.92 (0.87,0.97) | 0.44 | 0.91 (0.86,0.96) | 0.95 (0.87,1.03) | 0.40 |
| Folate | 100 µg | 0.84 (0.76,0.93) | 0.93 (0.83,1.04) | 0.19 | 0.85 (0.76,0.96) | 0.89 (0.80,0.99) | 0.67 | 0.98 (0.83,1.15) | 0.86 (0.79,0.94) | 0.17 | 0.88 (0.80,0.96) | 0.89 (0.78,1.03) | 0.89 |
| Carbohydrates | 50 g | 0.85 (0.78,0.94) | 0.93 (0.84,1.03) | 0.20 | 0.94 (0.84,1.04) | 0.85 (0.77,0.94) | 0.18 | 0.97 (0.83,1.12) | 0.88 (0.81,0.95) | 0.26 | 0.91 (0.83,0.99) | 0.86 (0.76,0.97) | 0.46 |
| Total sugars | 50 g | 0.87 (0.77,0.97) | 0.89 (0.79,1.00) | 0.79 | 0.92 (0.82,1.04) | 0.84 (0.75,0.95) | 0.29 | 1.01 (0.85,1.20) | 0.85 (0.77,0.93) | 0.09 | 0.91 (0.83,1.01) | 0.80 (0.69,0.93) | 0.16 |
| Red/proc. meat | 30 g | 1.08 (1.01,1.14) | 1.08 (1.01,1.16) | 1.0 | 1.06 (0.99,1.13) | 1.10 (1.03,1.17) | 0.43 | 1.10 (1.00,1.21) | 1.07 (1.01,1.12) | 0.62 | 1.06 (1.01,1.12) | 1.11 (1.02,1.20) | 0.35 |
| Fruit | 200 g | 0.90 (0.83,0.99) | 0.89 (0.81,0.98) | 0.87 | 0.88 (0.79,0.97) | 0.92 (0.84,1.01) | 0.53 | 0.98 (0.86,1.13) | 0.88 (0.82,0.95) | 0.17 | 0.90 (0.83,0.97) | 0.90 (0.80,1.02) | 1.0 |
| Vitamin C | 100 mg | 0.92 (0.83,1.02) | 0.87 (0.78,0.97) | 0.47 | 0.86 (0.77,0.96) | 0.93 (0.84,1.03) | 0.31 | 0.99 (0.85,1.15) | 0.87 (0.80,0.94) | 0.14 | 0.88 (0.81,0.96) | 0.92 (0.80,1.05) | 0.59 |
| Cereal | 40 g | 0.94 (0.88,1.01) | 0.91 (0.85,0.98) | 0.52 | 0.92 (0.86,0.99) | 0.94 (0.88,1.01) | 0.67 | 0.91 (0.83,1.01) | 0.94 (0.89,0.99) | 0.57 | 0.93 (0.88,0.99) | 0.93 (0.85,1.01) | 1.0 |
| Fibre | 5 g | 0.91 (0.84,0.99) | 0.92 (0.84,1.00) | 0.86 | 0.89 (0.81,0.97) | 0.92 (0.85,1.00) | 0.59 | 0.90 (0.80,1.03) | 0.92 (0.86,0.99) | 0.77 | 0.90 (0.84,0.97) | 0.94 (0.84,1.05) | 0.52 |
| Potassium | 1000 mg | 0.83 (0.74,0.94) | 0.95 (0.84,1.07) | 0.12 | 0.83 (0.73,0.94) | 0.90 (0.80,1.01) | 0.36 | 0.88 (0.74,1.05) | 0.90 (0.82,0.98) | 0.82 | 0.92 (0.83,1.01) | 0.82 (0.70,0.96) | 0.23 |

^1^Associations between each of the 17 foods or nutrients and colorectal cancer incidence separately using Cox proportional hazards regression models stratified by year of birth, date of completion of the 3-year re-survey (which is baseline for this study), and region of residence (10 geographical regions (9 in England), and Scotland), and adjusted for area-based deprivation (fifths, based on the Townsend deprivation score, unknown), highest educational qualification (none, technical, secondary, tertiary, unknown), body mass index (<20, 20-22.49, 22.5-24.9, 25.0-27.49, 27.5-29.9, 30-32.49, 32.5-34.9, 35+ kg/m2, unknown), height (<160, 160–164.9, ≥165 cm, unknown), strenuous exercise (none, ≤ once per week, > once per week, unknown), dietary energy intake (except for the analysis of energy and risk; fifths, unknown), alcohol (none, 1-5, 6-10, ≥ 11 drinks per week, unknown), smoking (never, past, current 1–4, current 5–9, current <10, current 10–14, current 15–19, current 20–24, current 25–29, current ≥30 cigarettes per day, unknown), and family history of bowel cancer (no, yes).
